# A multicenter Swedish histopathology image dataset of pediatric central nervous system tumors

**DOI:** 10.64898/2026.06.15.26355523

**Authors:** Per Nyman, Iulian Emil Tampu, Alia Shamikh, Gabriela Prochazka, Ida Blystad, Elisa Basmaci, Teresita Díaz de Ståhl, Pernilla Augustsson, Katarzyna Zielinska-Chomej, Duohui Cao, Jenny von Salomé, Arman Ardalan, Praveen Raj Somarajan, Gustaf Ljungman, Peter Lundberg, Johanna Sandgren, Neda Haj-Hosseini

## Abstract

Refined detection methods, more detailed tumor characterization, and adequate distinction between different pediatric tumor subtypes are necessary to improve diagnosis and treatment, enable precision medicine, and advance patient prognosis. However, the application of computational approaches to pediatric brain tumors remains limited, largely due to the lack of accessible datasets.

To address part of this gap, we provide whole slide images (WSIs) of hematoxylin and eosin (H&E)-stained tissue sections from pediatric central nervous system (CNS) samples collected in Sweden in 2013 to 2023. These data represent a population-based national cohort encompassing all six pediatric oncology centers in Sweden and are available through the Swedish Childhood Tumor Biobank (BTB). The dataset includes 1,446 WSIs of sufficient image quality with confirmed CNS tumor diagnoses, derived from 537 unique subjects (562 subject-diagnosis pair cases). In addition, diagnostic-relevant clinical information is included. Corresponding whole-genome sequencing (WGS), whole-transcriptome sequencing (WTS), and methylation array data are available upon application for most tumor samples through separate resources at BTB.

This H&E dataset has been specifically curated to support artificial intelligence-based analyses, while also serving broader applications in medical research and education. When combined with matched molecular data, it provides a valuable resource for advancing multimodal and precision diagnostic approaches in the pediatric population.

## Introduction

Tumors of the central nervous system (CNS) are the second most common type of cancer in children, after leukemia, and represent the most prevalent category of solid tumors in this population. There are numerous histological variants and biological subtypes of CNS tumors, reflecting their diverse nature. According to the WHO 2021 classification, these tumors are graded from 1 to 4. The most common pediatric CNS tumor, pilocytic astrocytoma, serves as an example of a low-grade glioma, typically associated with a favorable prognosis. In contrast, medulloblastomas and ependymomas are more aggressive pediatric tumors with poorer prognosis, often requiring intensive and prolonged treatment. Management of these tumors frequently involves chemotherapy and/or radiotherapy in addition to neurosurgery.

Preliminary diagnosis and evaluation during and after treatment are based on medical imaging, with magnetic resonance imaging (MRI) as the standard modality. The definitive diagnosis, however, is confirmed through histopathological examination and molecular analyses of tissue samples obtained during surgery. Traditionally, tissue sections are stained and investigated on glass slides under a microscope, but recently advances in digital pathology provide new opportunities for clinical examination through the sharing of scanned digital whole slide images (WSIs). In addition to histopathological review, routine clinical practice also includes molecular diagnostics, and with the development of digital pathology in recent years, artificial intelligence (AI) technology has also begun to be incorporated in tumor diagnostics. For instance, hematoxylin and eosin (H&E)-stained tumor sections, the routine histopathological staining technique for diagnostic evaluation, could benefit from AI-based analysis. When combined with molecular genetic data, computational approaches may enable more precise and accurate diagnostics, thereby supporting the advancement of precision and personalized medicine [1].

Most of the existing literature on the application of computational algorithms on histopathological brain tumor data has focused on adult gliomas [2]. In contrast, application of such approaches to pediatric brain tumors remains limited, primarily due to the scarcity of comprehensive pediatric histopathology datasets. Refined detection methods, more detailed tumor characterization, and adequate distinction between different subtypes of pediatric tumors are necessary to improve treatment strategies, support precision medicine, and improve patient outcomes. To enable the development of robust and reliable AI models, comprehensive datasets encompassing a broad spectrum of pediatric brain tumors are needed.

Because development of AI models typically requires large volumes of data, availability of multiple large datasets from diverse populations is highly beneficial. Together, such datasets can capture the broad spectrum of brain tumor types. Several resources already provide open-access WSI data for adult glioma, including TCGA/TCIA [3] [4], IvyGAP [6], CPTAC [7], U-PENN [8]. EBRAINS [9] extends this to additional diagnoses and includes H&E images from approximately 500 pediatric brain tumors. The Children’s Brain Tumor Network (CBTN) [10] [11] is currently the largest multimodal dataset for pediatric brain tumors, comprising over 2000 CNS tumor cases. Despite the availability of these resources, there remains a need for well-curated, AI-adapted datasets to support robust analysis and reliable model development in pediatric brain tumor research.

Joining the global efforts, we present a comprehensive description of a retrospectively collected dataset of H&E-stained WSIs, paired with clinical information from a robust national cohort. The cohort includes samples collected from all six pediatric oncology centers in Sweden, during the period 2013-2023, and is available upon application through the Swedish Childhood Tumor Biobank (Barntumörbanken, BTB). The dataset has been specifically adapted for AI-based analysis and constitutes a valuable resource for histopathological research, education, and benchmarking of computational methods. When integrated with molecular features derived from WGS and WTS, such as driver mutations, gene fusions, and copy-number alterations, it enables multimodal diagnostic approaches that reconcile morphology with genotype. This integration may improve and accelerate tumor classification, resolve diagnostically challenging cases, and support risk stratification and treatment selection.

## Data Collection

### Study Approval

Ethical approval was obtained from the Swedish Ethical Review Authority (Dnr 2021–03985,with amendments Dnr 2022-00065-02 and 2024-07014-02) permitting the conduct of this research study and access to relevant resources (H&E slides, WSI, and diagnostic data) from BTB. The approval also covered digitization of a subset of the glass slides, curation of the WSIs in Linköping, Sweden and AI-based analysis of the data. An application, including the ethical permit, was submitted to BTB at the Karolinska University Hospital (K) to obtain access to H&E slides, WSIs, and diagnostic data. K is the formal controller of BTB’s personal research data, while the Stockholm Medical Biobank (SMB) serves as the medico-legal authority for BTB’s tissue samples. Applications for both data and samples were approved by BTB, K, and SMB.

Pediatric patients with suspected solid tumors who are scheduled for biopsy/surgery, and/or their legal guardians, depending on the age of the child eligible for surgery, are asked to provide informed consent for inclusion in the BTB biobank project. BTB is an ongoing ethically approved research initiative involving sample-and data collection, as well as an established national infrastructure for childhood tumors. BTB methodically collects tissue samples and generates genomic and other omics-data from pediatric patients with solid intracranial and extracranial tumors to support and facilitate research in the field for future healthcare advances for affected children [12].

Multiple sample types and associated patient data, including H&E WSIs and diagnostic information, are continuously collected nationwide in Sweden in accordance with established biobank agreements. The primary biospecimens include fresh-frozen tumor tissue and whole-blood samples, although additional materials such as cerebrospinal fluid and viable tumor cells may also be collected. Samples and data are systematically and securely organized and stored at BTB. Access to biospecimens and associated datasets for secondary use is granted following a formal application procedure and requires separate ethical approval. Subject to consent and provided that sufficient material remains available after clinical diagnostics, slides and/or WSIs are prepared as part of clinical routine for storage in the biobank, along with relevant clinical diagnostic information for the purposes of the present study.

### Sample Collection and Data Overview

In addition to fresh frozen tissue, all six pediatric oncology centers in Sweden periodically deliver formalin-fixed paraffin-embedded (FFPE) H&E-stained tissue samples to BTB, either as glass slides or as digitized WSIs. The H&E-stained sections are prepared within the routine clinical diagnostic procedure. The sample collection presented here includes all glass slides and WSIs supplied within the period 2013 – 2023 relating to CNS tumors. WSIs from Linköping, Uppsala and Umeå as well as glass slides from Lund, Gothenburg and Stockholm were obtained from BTB and further scanned in Linköping as part of this project, aiming to create a retrospectively collected unified digital pathology dataset Figure 1.

**Figure 1:**
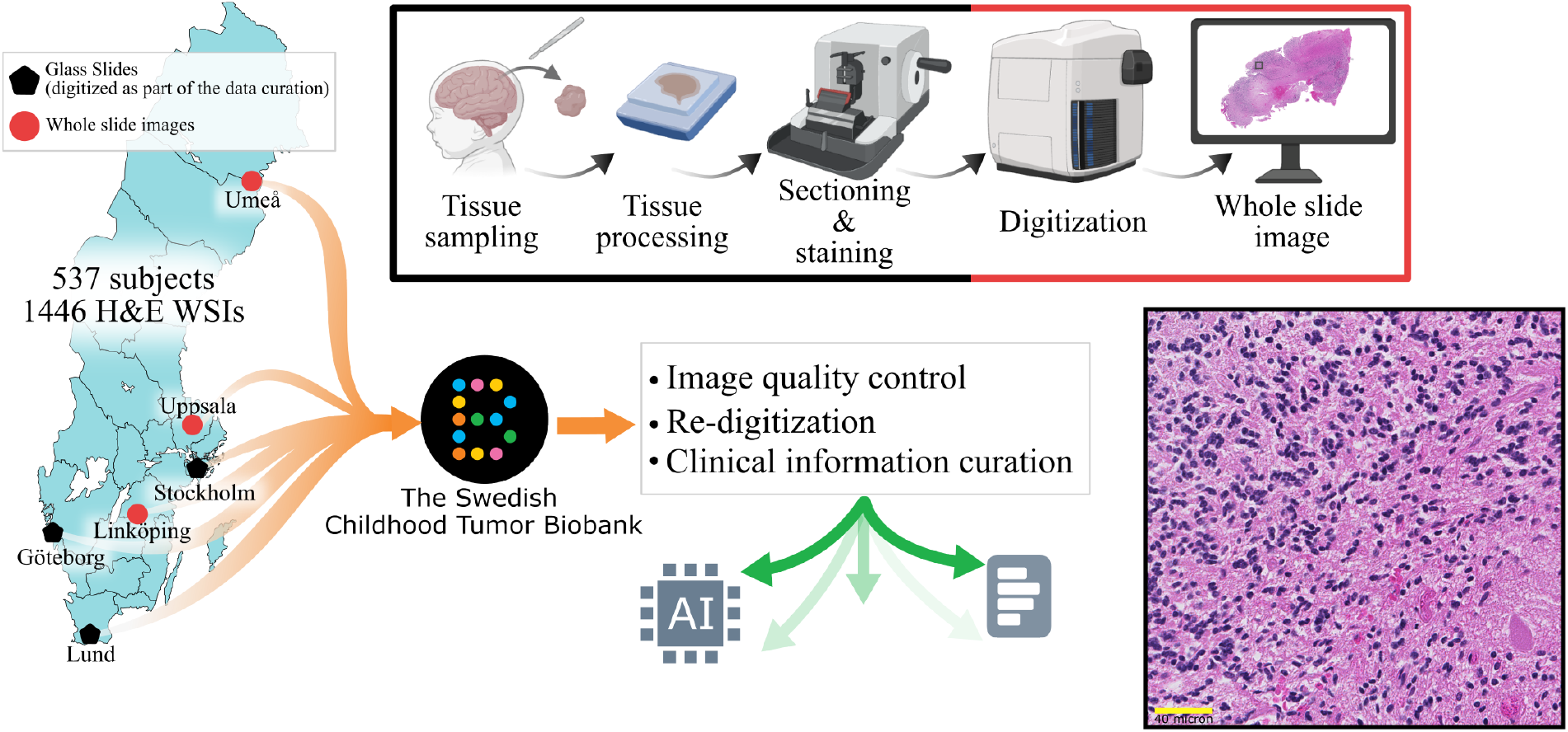
Illustration of the dataset production process, including collection, scanning and curation (Created with BioRender.com).

The total number of slides from all six centers was 1446, derived from 562 cases and 537 subjects. Cases are defined as subject-diagnosis pairs, including relapses and metastases associated with the primary diagnosis. Following diagnosis matching, image quality control, and clinical information curation, a total of 516 subjects remained in the dataset (Figure 2). Considering only the unique subjects with available clinical information, acceptable image quality, confirmed CNS localization, and verified CNS tumor diagnosis, the final cohort comprised 267 males and 232 females. The age range of the subjects was 0-18 years; additionally, three subjects (six cases) aged 19-25 years were included due to recurrence following an initial diagnosis before age 18. For one subject, age information was unavailable. The age and sex distribution of the subjects (based on primary cases only) are illustrated in Figure 3.

**Figure 2:**
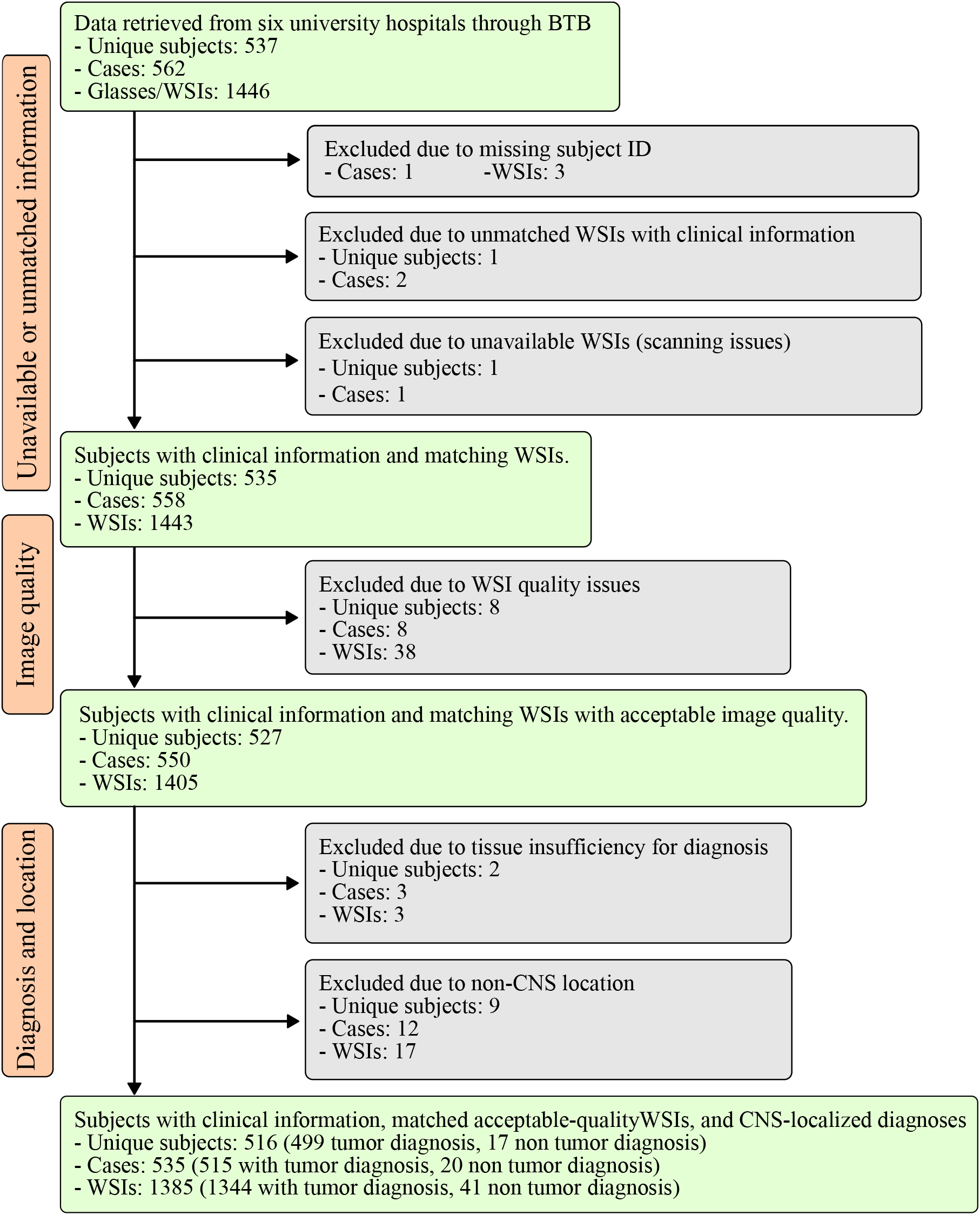
Data inclusion and exclusion flow diagram. Green boxes indicate the available data, while the gray and orange boxes on the left side represent the number of exclusions and the reasons for exclusion, respectively. The bottom box shows the final CNS dataset after all exclusions.

**Figure 3:**
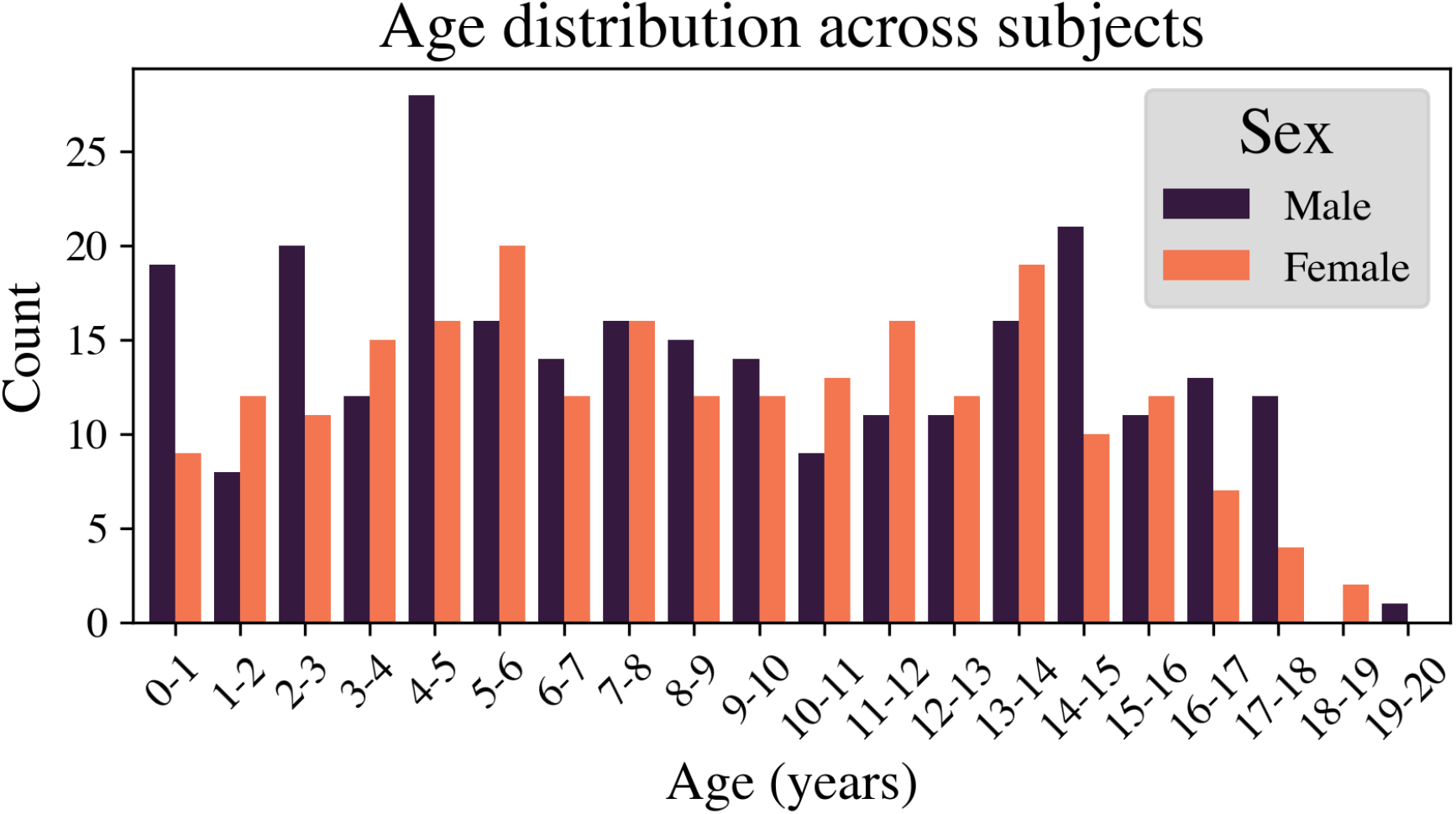
Age distribution by sex across subjects with CNS tumors after exclusions (267 males and 232 females). In the 18 cases where scans were available for both primary disease and recurrence, only the age at primary diagnosis was included in the distribution.

### WSI Quality Control

Before scanning, the glass slides from Stockholm, Gothenburg and Lund were cleaned with 80% alcohol to remove dust and erase pen markings. Scanning was performed using a Hamamatsu_Nanozoomer XR & S360-scanner (Hamamatsu Photonics K.K., Japan) at 40*x* magnification. The resulting images were saved in ndpi format. After completion of scanning, the glass slides were returned to BTB.

All the WSIs were manually reviewed for scan quality by two independent readers(non-pathologists). Scans that did not meet the predefined quality criteria were re-scanned when corresponding glass slides were available. The criteria for passing the quality assessment were:

1. all tissue regions present on the glass slide were included in the scan.
2. all the tissue regions were in focus at 20× magnification, including areas affected by glass scratches or other glass-related artifacts, as well as blood-containing region.
3. the WSI did not show excessive dust or pen markings over the tissue regions.
4. the WSI contained tissue.

A second quality assessment was then performed on the re-scanned WSIs, with those that failed to meet the quality criteria subjected to a third and final scan. For the WSIs that still failed to meet all of the criteria after the third scan, the WSI with the higher quality was included. Of the available 543 glass slides, four were not scanned due to either earlier slide breakage (n=3) or absence of tissue (n=1). Figure 4 shows examples of artifacts considered during the quality assessment as well as a representative WSI with no artifacts.

**Figure 4:**
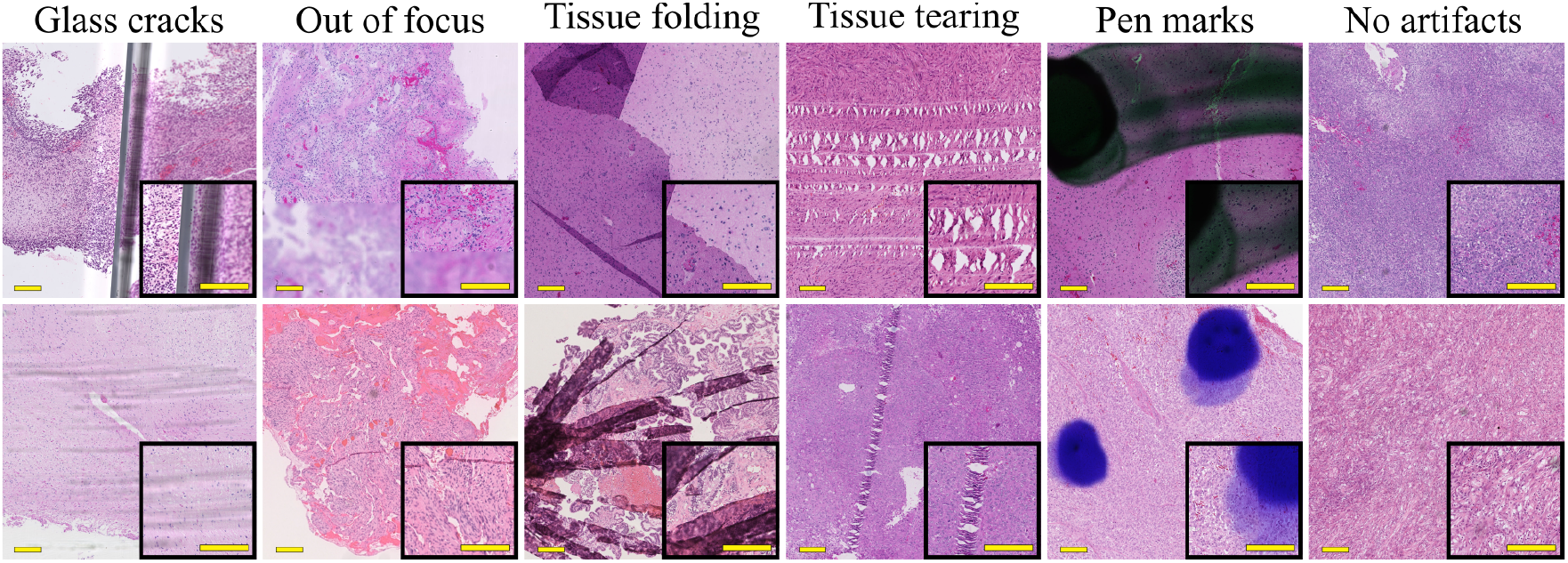
Examples of WSI scans illustrating representative artifacts. Overview images were acquired at 5× magnification, with insets highlighting selected areas at 20× magnification. Scale bars correspond to 200µm in the overview images and 100µm in the insets.

WSIs from Linköping, Umeå, Uppsala, that were delivered as images (in ndpi or svs format) from the clinical routine, were evaluated using the same quality criteria as above by the same two independent readers. For cases that did not meet the criteria, glass slides were requested from the original centers for a second review and rescanning. Finally, WSIs (n=38) that did not meet the quality criteria were excluded from the final dataset, leading to exclusion of 8 subjects without any remaining WSIs. A summary of the participating centers, the number of WSIs and subjects, scanner specifications, and scanning magnification (20× or 40×) and the final counts are provided in Table 1.

**Table 1:** Overview of the data collected from the contributing centers. Linköping, Umeå and Uppsala provided data as images scanned in the clinical routine.

| Site | Scanner/magnification<br>(File format) | No. of subjects<br>(% over entire dataset) | No. of WSIs<br>(% over entire dataset) |
| --- | --- | --- | --- |
| Stockholm | Hamamatsu Nanozoomer_XR & S360<br>40× (.ndpi) | 139<br>(25.9%) | 230<br>(15.9%) |
| Gothenburg | Hamamatsu Nanozoomer_XR & S360<br>40× (.ndpi) | 96<br>(17.9%) | 294<br>(20.3%) |
| Lund | Hamamatsu Nanozoomer_XR & S360<br>40× (.ndpi) | 71<br>(13.2%) | 128<br>(28.8%) |
| Linköping | Hamamatsu Nanozoomer<br>20× (.ndpi, .svs) | 63<br>(11.7%) | 212<br>(14.7%) |
| Umeå | Hamamatsu Nanozoomer S360 40× (.ndpi)<br>40× (.ndpi) | 38<br>(7.1%) | 182<br>(12.6%) |
| Uppsala | Hamamatsu Nanozoomer S60<br>20× (.ndpi, .svs) | 130<br>(24.2%) | 400<br>(27.7%) |
| Total |  | 537 | 1446 |

**Table 2:** Description of the curated data records and clinical (WHO2021) corresponding to the images.

| Variable | Description of clinical information |
| --- | --- |
| Center | Stockholm, Gothenburg, Lund, Linköping, Umeå, Uppsala |
| Subject ID | Alphanumeric identifier |
| WSI ID | Alphanumeric identifier |
| Sex | Male/Female |
| Age at diagnosis | In months |
| Diagnosis | Not tumor, Not malignant<br>Malignant (tumor category/ family/ type /subtype) |
| Localization | Infratentorial, Supratentorial, Spinal<br>Unknown, Extradural, Meninges, Not CNS |

### Clinical Information and Curation

In this dataset, 17 subjects and 20 cases (41 WSIs) were non-tumors and instead presented a final diagnosis consistent with inflammation, hemorrhage, gliosis, and unspecified tissue anomalies. A total of 516 cases had a confirmed tumor diagnosis. Based on the WHO 2021 classification guidelines, the dataset encompasses 10 tumor categories, 26 tumor families, and 39 tumor types (Figure 5 and Table S1 in the supplementary material). It should be noted that these tumor categories, families, and types reflect only the entities represented in this dataset and do not cover all the entities available in the WHO 2021 guideline. One subject (one case of high-grade neoplasm without further specification) could not be classified according to WHO2021 criteria. Additional clinical variables, patient sex and age in months at diagnosis and tumor localization in CNS (infratentorial/ supratentorial/ spinal/extradural/ meninges/Unknown) were also recorded. Distribution of the tumor localizations is illustrated in Figure 6. The clinical information is available as a tabular file. This dataset is prepared in accordance with FAIR principles [13] and is adapted to support AI-based analysis.

**Figure 5:**
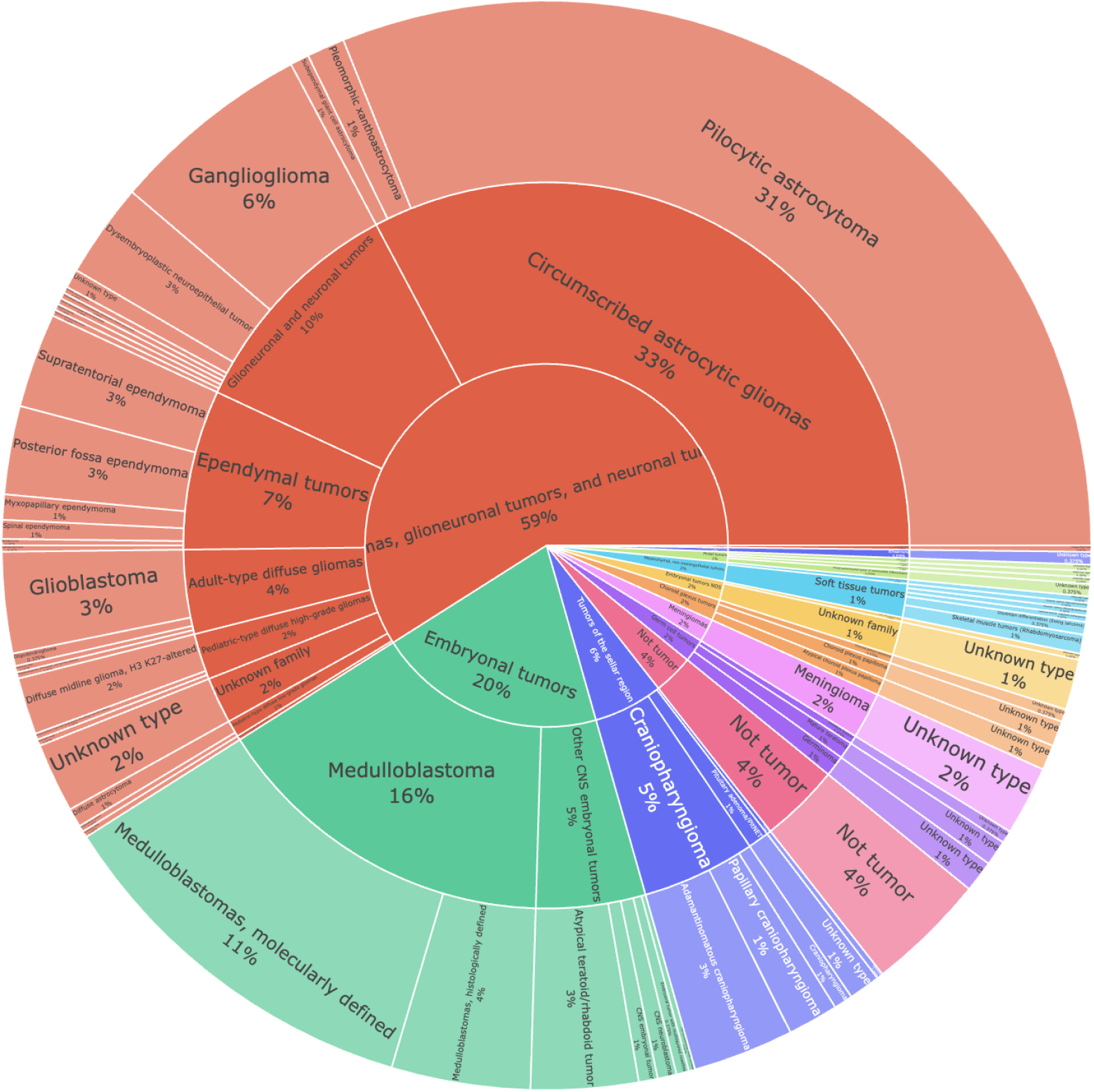
Hierarchical tumor classification distribution (subject count) in the final dataset according to diagnosis presented by tumor category, family and type. Details of the information can be found in Table S1 in the Supplementary Material. [See the interactive Figure](https://medical-image-analysis-group-at-liu.github.io/BTB_dataset_descriptor/).

**Figure 6:**
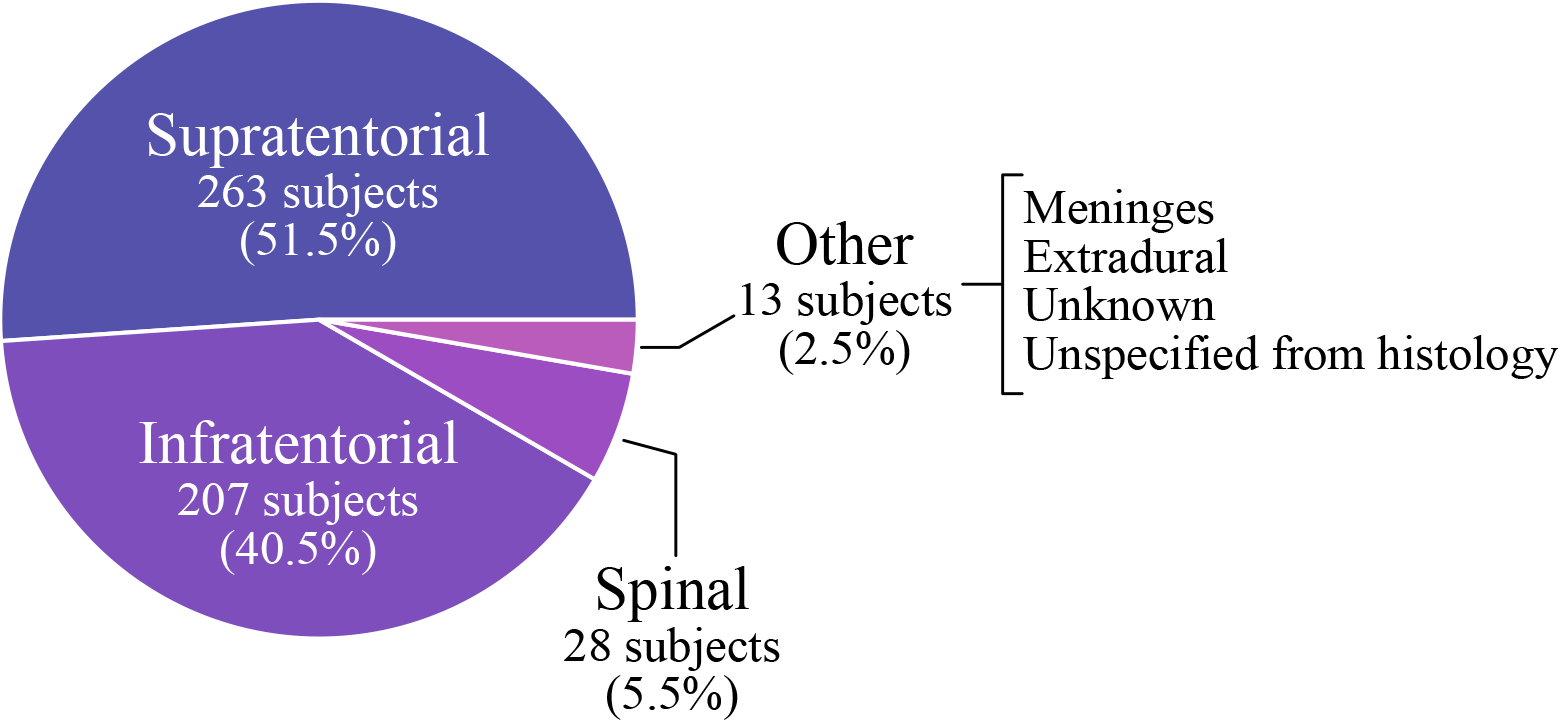
CNS Tumor distribution by location based on the curated clinical information. The figure reflects the final cohort described in the last box of Figure 2. The total number of subjects appears higher when summed across locations, as some subjects present tumors in multiple anatomical locations.

## Technical Validation

The technical validation of this dataset is described in [14] where WSIs are used to predict the diagnoses, in terms of tumor category, family and type, using deep learning methods. Specifically, the UNI and CONCH foundation models [15][16] were used as feature extractors under a multiple instance learning framework. Furthermore, the methodology was evaluated on an additional pediatric brain tumor dataset (CBTN) fused with immunohistochemistry data, demonstrating that the integration of H&E and IHC WSIs improves the predictions [17]. As reported in [14] recent computational pathology foundation models are considerably less affected by the artifacts such as glass cracks and pen marks compared to the earlier models, thereby reducing the need for augmentation and preprocessing steps, e.g., stain normalization or artifact removal. In addition, the technical validation highlights that tissue area on the slides, alongside the number of WSIs, represents an important quality parameter in data preparation that affects model performance.

### Summary

A dataset of histology WSIs from all the pediatric oncology centers in Sweden was curated and organized into both image and tabular formats, suitable for AI-based analysis, as well as conventional scientific research. AI technologies have the potential to become valuable decision-support tools, enabling clinical implementation and ultimately contributing to more personalized treatment strategies in CNS tumor diagnostics for children and adolescents. Development of such methods requires large volumes of data, and access to multiple datasets from diverse populations covering the full spectrum of brain tumor types, is essential for training and validating neural network models for automatic tissue classification based on the WSIs.

The data collection will continue over time to incorporate additional samples, and the dataset will be updated accordingly. Molecular and genetic information, whole-genome sequencing, whole-transcriptome analysis, and methylation profiling matched to the WSIs, are currently available as a separate dataset [12]. Future work aims to integrate MR imaging, WSIs, genetic information, and detailed clinical data to support the development of comprehensive AI-based decision support systems for image diagnostics.

## Data Availability

Instructions for obtaining data and sample images from BTB are available upon formal request at www.barntumorbanken.se. Access requires appropriate ethical approval, compliance with BTB requirements, and submission of an application that undergoes a review process. Upon approval, data access agreements are established prior to data release.

## Author contribution

All authors reviewed and approved the final manuscript.

## Acknowledgments

The authors gratefully acknowledge The Swedish Childhood Tumor Biobank, supported by the Swedish Childhood Fund for access to samples and data. The study was financed by Swedish Childhood Cancer Fund (MT2021-0011, MT2022-0013 (NHH), and MT2019-0027, PR2020-0071 (PL)), Joanna Cocozza’s Foundation for Children’s Medical Research (2023 and 2025 (NHH)); Linköping University’s Cancer Strength Area (2022, 2024), Vinnova project 2017 02447 via Medtech4Health and Analytic Imaging Diagnostics Arena (AIDA, 1908) (NH) and ALF Grants, Region Östergötland (974566) (IB).

The valuable assistance of the clinical pathology departments and pediatric oncology centers at University Hospitals in Gothenburg, Linköping, Lund, Stockholm, Uppsala and Umeå is gratefully acknowledged. Use of BioRender.com for preparation of parts of Figure 1 is acknowledged under license (BG29TSUOFY).

## Ethics Statement and Patient Consent

Ethical approval was obtained from the Swedish Ethical Review Authority (Dnr 2021–03985, with amendments Dnr 2022-00065-02 and and 2024-07014-02). The study was additionally approved by BTB, as well as Karolinska University Hospital and Stockholm Medical Biobank, the medico-legal authority for BTB’s personal data and tissue samples, respectively.

## Supplementary Material

**Table S1:**
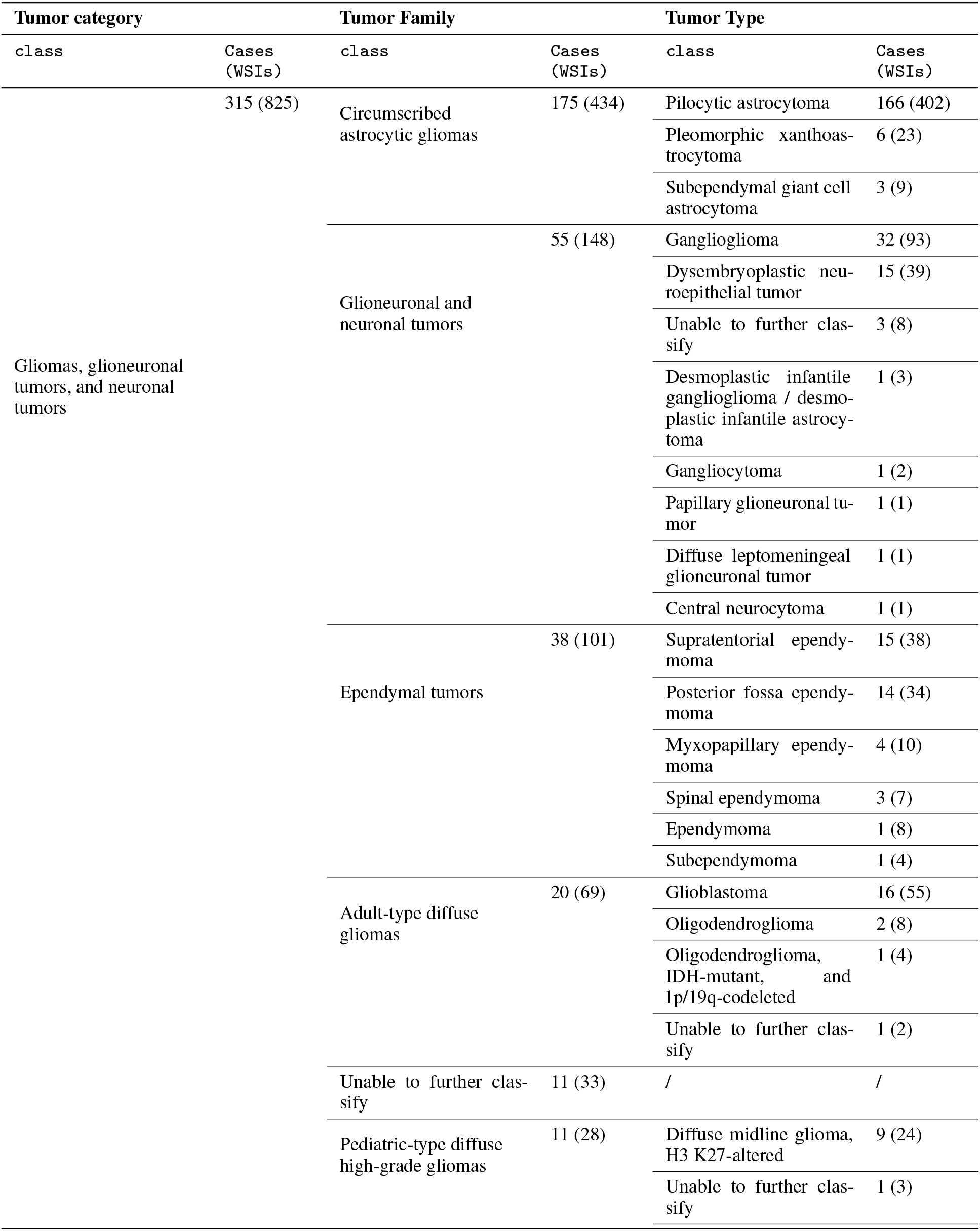
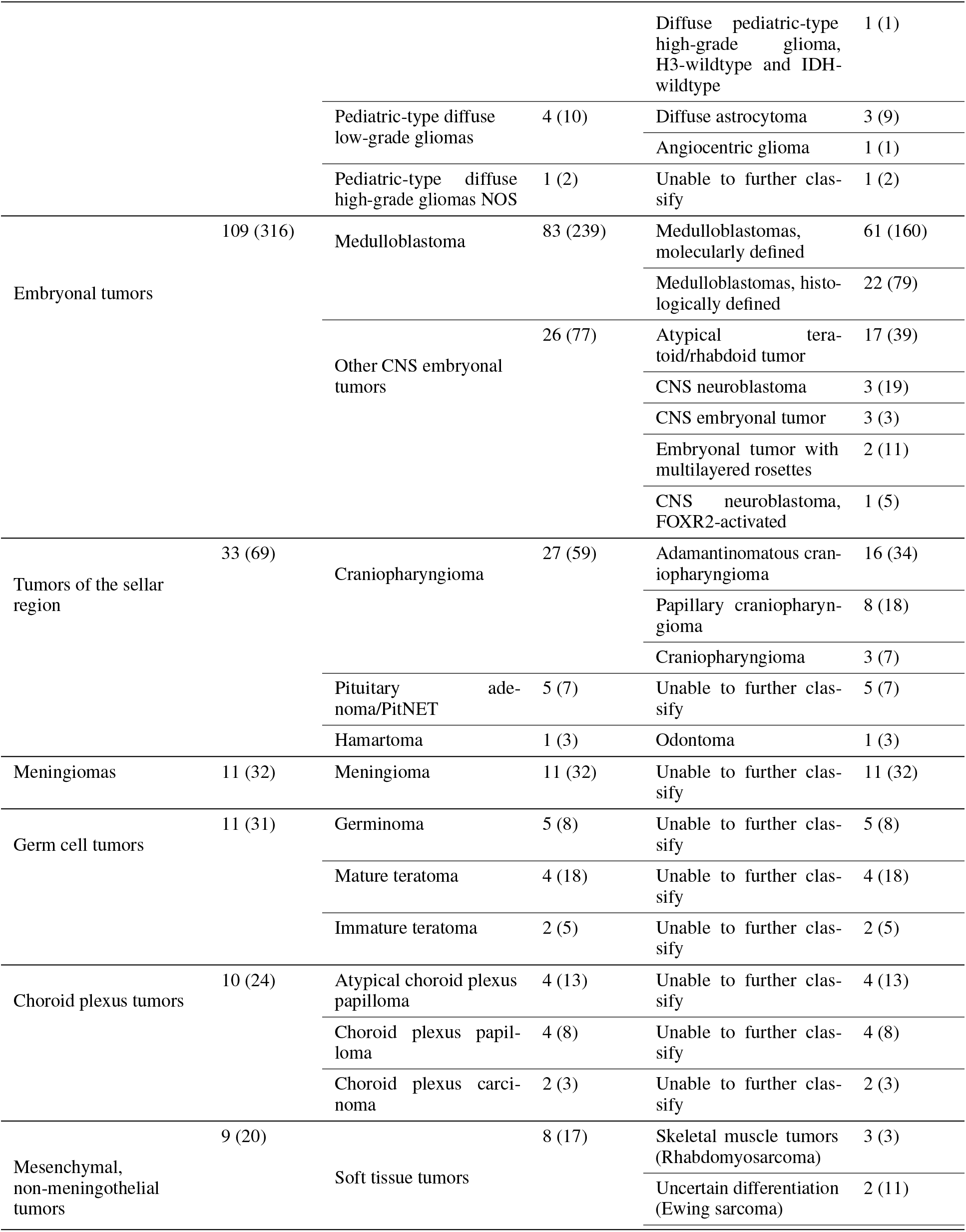
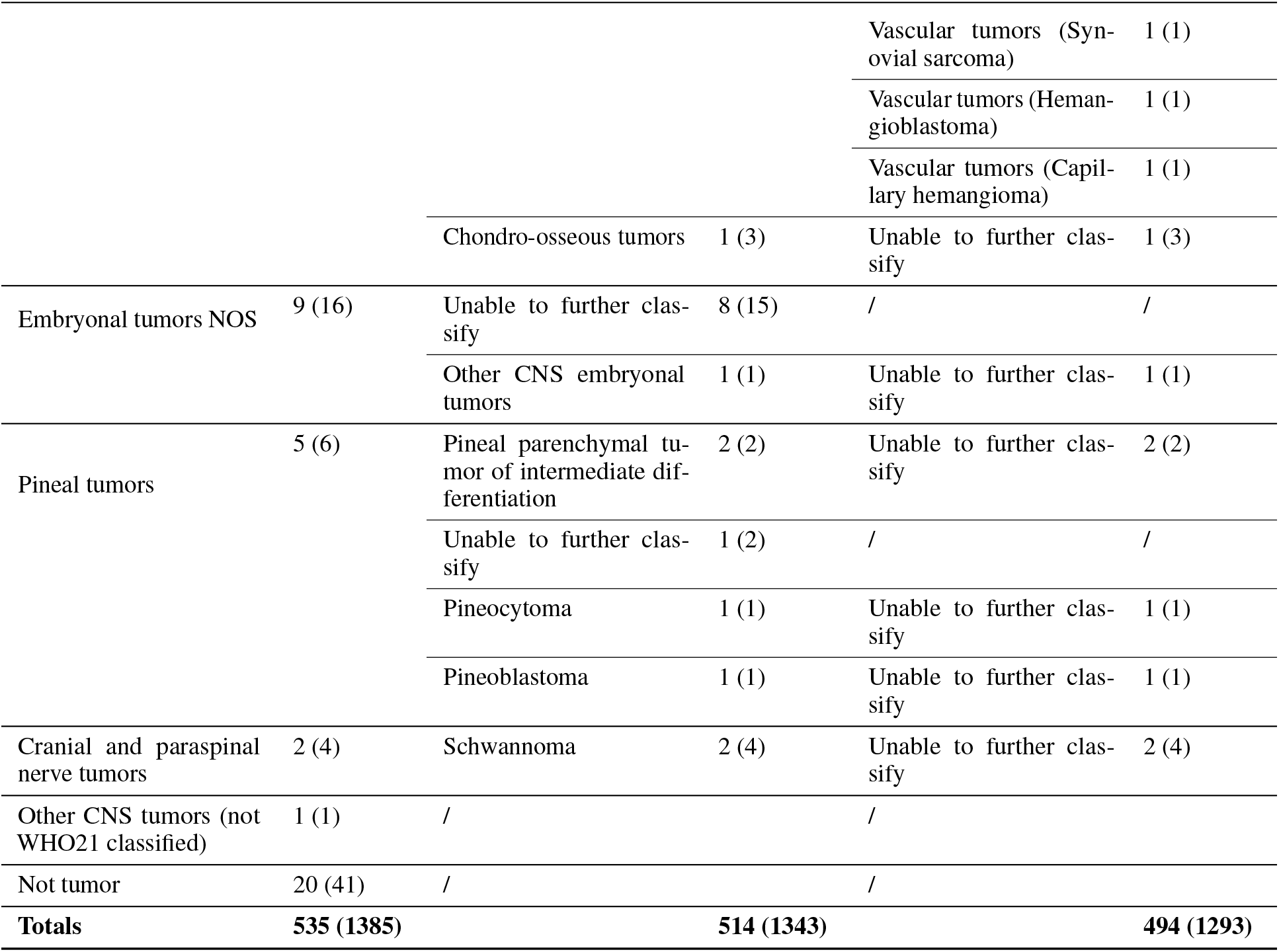
Tumor classification overview by category, family, and type. Case counts and WSI counts are provided for each level.

## References

[1] D. N. Louis et al., “The 2021 who classification of tumors of the central nervous system: A summary,” Neurooncology, vol. 23, no. 8, pp. 1231–1251, 2021.

[2] J.-P. Redlich et al., “Applications of artificial intelligence in the analysis of histopathology images of gliomas: A review,” npj Imaging, vol. 2, no. 1, p. 16, 2024.

[3] K. Clark et al., “The cancer imaging archive (tcia): Maintaining and operating a public information repository,” Journal of digital imaging, vol. 26, no. 6, pp. 1045–1057, 2013.

[4] “The cancer genome atlas program,” Accessed: 2024. [Online]. Available: https://www.cancer.gov/ccg/research/genome-sequencing/tcga.

[5] R. B. Puchalski et al., “An anatomic transcriptional atlas of human glioblastoma,” Science, vol. 360, no. 6389, pp. 660–663, 2018.

[6] “Allen institute’s ivygap data.” Accessed: 2021. [Online]. Available: http://glioblastoma.alleninstitute.org/.

[7] “The clinical proteomic tumor analysis consortium glioblastoma multiforme collection (cptac-gbm) (version 16) [dataset],” Accessed: 2024. [Online]. Available: 10.7937/K9/TCIA.2018.3RJE41Q1.

[8] S. Bakas et al., “The university of pennsylvania glioblastoma (upenn-gbm) cohort: Advanced mri, clinical, genomics, & radiomics,” Scientific data, vol. 9, no. 1, p. 453, 2022.

[9] T. Roetzer-Pejrimovsky et al., “The digital brain tumour atlas, an open histopathology resource,” Scientific Data, vol. 9, no. 1, p. 55, 2022.

[10] “Children’s brain tumor network (cbtn),” Accessed: 2023. [Online]. Available: https://cbtn.org/.

[11] J. V. Lilly et al., “The children’s brain tumor network (cbtn)-accelerating research in pediatric central nervous system tumors through collaboration and open science,” Neoplasia, vol. 35, p. 100 846, 2023.

[12] T. Díaz de Ståhl et al., “The swedish childhood tumor biobank: Systematic collection and molecular characterization of all pediatric cns and other solid tumors in sweden,” Journal of Translational Medicine, vol. 21, no. 1, p. 342, 2023.

[13] M. D. Wilkinson et al., “The fair guiding principles for scientific data management and stewardship,” Scientific data, vol. 3, no. 1, pp. 1–9, 2016.

[14] I. E. Tampu et al., “Pediatric brain tumor classification using digital pathology and deep learning: Evaluation of sota methods on a multi-center swedish cohort,” Brain Pathology, vol. 36, no. 1, e0029, 2026. DOI: 10.1111/bpa.70029.

[15] M. Y. Lu et al., “A visual-language foundation model for computational pathology,” Nature medicine, vol. 30, no. 3, pp. 863–874, 2024.

[16] R. J. Chen et al., “Towards a general-purpose foundation model for computational pathology,” Nature medicine, vol. 30, no. 3, pp. 850–862, 2024.

[17] C. Spyretos, I. E. Tampu, J. Lindblad, and N. Haj-Hosseini, “Multi-stain fusion of histopathology images using deep learning for pediatric brain tumor classification,” bioRxiv, pp. 2026–04, 2026.

